# Private Choices in Public Health: Endogenous Behavioral Responses to Pandemic Risk

**DOI:** 10.1101/2025.11.08.25339826

**Authors:** J. Felipe Montano-Campos, Kyueun Lee, Blythe Adamson, Anirban Basu

**Author notes:** **Corresponding Author**: J. Felipe Montano-Campos, PhD.

## Abstract

Epidemic control depends not only on policy mandates but also on endogenous behavioral responses—how individuals adjust private preventive actions to evolving risk. Using over 1.1 million county-day observations from 1,206 U.S. counties, we integrate high-frequency mobility data with local COVID-19 mortality to estimate the semi-elasticity of preventive behavior to mortality-driven risk perception. Fixed-effects regressions exploit within-county temporal variation, controlling for day-of-week and month effects, county-specific linear trends, and public health policies. Behavioral elasticity evolved as the pandemic progressed. Early in the pandemic, mortality-driven risk perception had only a limited effect on behavior (3.1% reduction in mobility per additional death per 10,000 residents, 7-day lag), while shelter-in-place mandates drove large declines (–10.16%). As uncertainty declined and individuals learned about transmission risks, behavioral responsiveness strengthened markedly, with mobility falling 7.8% per additional death (7-day lag per 10,000)—indicating that risk-calibrated adaptation supplanted broad precautionary withdrawal. By the post-vaccination stage, the mortality–mobility association was no longer significant, consistent with behavioral desensitization as perceived risk fell. To unpack the selective logic of prevention—when behavior most closely tracked perceived risk—we find that reductions were most pronounced in discretionary settings (–5.2% retail/recreation; –5.3% transit) but statistically insignificant in essential activities (–0.4% grocery/pharmacy) and low-risk outdoor spaces (–1.7% parks). Responsiveness weakened by roughly 0.5% per month, while shelter-in-place and mask mandates amplified mortality-driven behavioral responses by 2.8% and 4.7%. These findings highlight that voluntary, risk-responsive behavior—amplified by policy interventions—was pivotal to epidemic control.

## Introduction

In public health crises, individual behavior is not merely reactive—it is endogenous, as it is shaped by perceived risk, which in turn evolves with epidemic outcomes driven by past behavior. Understanding how individuals adjust their private preventive behaviors and voluntary actions to reduce infection risk in response to evolving health risks is critical for managing infectious disease outbreaks and designing effective public health policies. The COVID-19 pandemic represents an unprecedented natural experiment, exposing communities worldwide to rapidly changing and obvious health risks, thus providing a unique opportunity to study endogenous dynamic behavioral responses to risk signals empirically. The economic principle of risk compensation is central to understanding these responses, describing how individuals adjust their private preventive behaviors —voluntary actions undertaken to mitigate the risk of infection— in response to perceived risk. In this paper, we study how local observed mortality signals influence community private preventive behaviors for COVID-19, specifically reductions in mobility, presumably through forming individual risk perceptions. Our central focus is on endogenous behavioral responses that operate alongside, and sometimes independently from, formal public health policies. The presence of such private endogenous behavior has long been theorized in the economics literature [1]; however, it has not been comprehensively studied empirically in the context of COVID-19.

Risk compensation is critical for accurately evaluating public health policies and intervention effectiveness [2], [3], [4], [5]. The success of such policies hinges not only on their direct effects but on how individuals adapt their behavior in response. Yet policy design often overlooks the endogenous relationship between perceived risk and private behavior—treating responses as fixed rather than adaptive. As articulated by the Lucas critique, once a policy is implemented, individuals rapidly adjust their beliefs, expectations, and behaviors, resulting in unforeseen outcomes. Traditional epidemiological models deployed during the COVID-19 times often assumed static or constant transmission rates, neglecting the dynamic nature of human behavior and risk perception adjustments [6], [7], [8]. This oversight can cause serious misestimations of policy effectiveness. Models may overstate long-term impact if they ignore declining demand for prevention as perceived risk falls, or understate it if they assume uniform rather than heterogeneous responses across diverse populations. Such simplifications overlook key behavioral dynamics, weakening policy outcomes and public health strategies. For example, COVID-19 shelter-in-place orders initially led to significant mobility reductions, but compliance waned over time as risk perceptions adjusted [9], [10]. Neglecting these behavioral adaptations can undermine policy effectiveness, prolong outbreaks, and elevate risks, underscoring the need to study how individuals adjust to evolving signals to design robust interventions.

Previous evidence across multiple health interventions consistently highlights the presence of risk compensation behaviors. In infectious disease prevention, in the context of HIV, major treatment breakthroughs and the introduction of pre-exposure prophylaxis (PrEP) led to increased sexual risk-taking, including higher rates of condomless sex and STI incidence, as improved survival expectations reduced perceived risk [11] [12], [13] [14]. Similar behavioral adjustments have been documented during the COVID-19 pandemic. Following vaccination or mask mandates, individuals reported lower adherence to protective behaviors such as masking, distancing, or home confinement, and increased mobility in public spaces [15][16][17].

However, these studies primarily focus on behavioral shifts following the adoption of specific protective measures, offering limited insight into how individuals adjust their behavior in response to evolving epidemic dynamics. Our study offers a novel perspective: instead of looking at behavior after protection is adopted, we examine how individuals proactively adjust preventive behaviors in response to real-time, objective risk signals — specifically, COVID-19 mortality. We focus on mortality, which offers one of the most salient and behaviorally influential signal during outbreaks. Unlike case counts, which are distorted by limited testing access, underreporting, and delays, deaths offer a stable, visible, and emotionally resonant measure [18], [19]. Individuals respond more strongly to concrete outcomes like death, especially when amplified by media [20], [21], making mortality a powerful anchor of perceived threat [22].

We used daily county-level data on COVID-19 cases, deaths, and mobility across six domains to capture multiple dimensions of private preventive behavior, allowing for a nuanced characterization of risk compensation dynamics. We then traced how these behaviors evolved as the pandemic progressed and examined how public health policies—specifically shelter-in-place orders and mask mandates—modified these adjustments, clarifying the dynamic interplay between perceived risk and policy interventions.

### Study Timeframe

Our study spans from February 15, 2020, to November 30, 2021, covering key phases of the COVID-19 pandemic in the United States. This timeframe ensures that we capture the evolution of risk perception and preventive behaviors during the pandemic while maintaining a clean analytical period that allows us to assess behavioral responses under a period where vaccines were highly effective before later waves introduced complexities related to waning immunity and variant resistance (Omicron variant).

### Data Sources and Variable Construction

#### COVID-19 Cases and Deaths

To track the progression of COVID-19 across the United States, we used daily county-level confirmed case and death counts from the Johns Hopkins Coronavirus Resource Center (JHU CRC). The JHU CRC became the primary global repository for real-time infectious disease tracking, aggregating data from 260 sources, including 182 local, state, and federal agencies, such as the Centers for Disease Control and Prevention (CDC), the World Health Organization (WHO), and state health departments [23]. We apply a seven-day moving average to daily case and mortality counts to smooth short-term fluctuations and reporting inconsistencies, thereby stabilizing trends and improving estimate accuracy.

#### Mobility Data

We developed a novel measure of domain-specific absolute mobility by integrating two complementary high-frequency data sources: SafeGraph’s absolute mobility levels and Google’s category-specific mobility trends. Whereas Google data alone provide only relative percentage changes from a pre-pandemic baseline, our integrated approach reconstructs absolute, county-day mobility across six domains—Retail & Recreation, Grocery & Pharmacy, Workplaces, Transit Stations, Parks, and Residential. This allows us to capture the intensity of movement by activity type, enabling fine-grained analysis of how individuals adjusted preventive behaviors in response to evolving risk and policy environments. (See Appendix for details on construction, calibration, and validation.)

#### Public Health Policy Measures

To capture the influence of public health interventions on private preventive behaviors, we included two policy variables: Shelter-in-Place (SIP) Orders and Mask Mandates. Data came from the U.S. Department of Health & Human Services’ COVID-19 State and County Policy Orders dataset, which compiles daily county-level records of mandates nationwide. We defined binary indicators for whether a SIP order or mask mandate was active on a given day in a given county. SIP orders restricted movement except for essential needs, while mask mandates required face coverings in public settings.

### Empirical Strategy

#### Pandemic Period Classification

Given the evolving nature of the pandemic, we divided our analysis into four stages, reflecting changes in public knowledge about the virus, levels of uncertainty, perceived risk, and patterns of individual behavioral response—defined by contextual shifts rather than fixed epidemiological thresholds. The Early Pandemic (Feb–Apr 2020) Period was marked by fear and uncertainty, with private preventive behaviors driven more by generalized threat perception than informed self-protection. The Risk Compensation Period (May 2020–Feb 2021) reflected a shift toward more calibrated decisions, as individuals weighed trade-offs and adjusted behavior in response to local mortality signals. The Vaccination Rollout/Transitional Period (Mar–May 2021) was a transitional stage with mixed conditions. And, the Post-Vaccination Period (Jun–Nov 2021), declining perceived threat and confidence in immunity shaped behavioral choices, reducing the salience of mortality driven risk perception as a driver of private prevention. (See Appendix for detailed definitions and criteria.)

#### Empirical Model and Estimation Strategy

Our analysis assumes that observed mortality shapes community behavior by influencing perceived risk. While we do not observe perception directly, we interpret behavioral responses to local mortality as operating through this perception channel. We restricted the sample to counties with at least 30 days of mobility and mortality data during the Risk Compensation period to ensure stable estimates and reduce noise from sparse reporting.

We use mobility as a proxy for private preventive behavior, with reductions reflecting voluntary efforts to lower infection risk by avoiding public spaces. Our main outcome variable is total mobility, aggregating movement in Retail & Recreation, Grocery & Pharmacy, Workplaces, and Transit Stations, while excluding parks since outdoor activity reflects safer substitution rather than true risk avoidance. Excluding park mobility ensures that our measure of private preventive behavior is more directly tied to risk-avoidance behaviors in high-exposure settings. To formally examine the relationship between risk perception and private preventive behavior, we estimated the following baseline fixed effect regression model

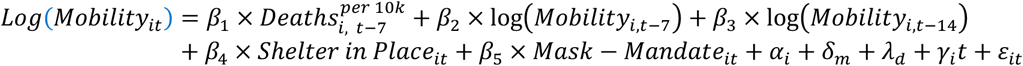

Our main empirical specification uses a log-linear regression model to estimate the relationship between COVID-19 mortality risk perception and mobility behavior. The dependent variable, *Log*(*Mobility_it_*), represents the log-transformed daily mobility level in county *i* on day *t*, normalized by the county population to account for differences in population size across regions. The key independent variable, *Deaths_i, t-7_*, measures the number of deaths per 10,000 people in county *i* lagged by seven days, capturing how mortality information from the past week influences current mobility decisions. We lagged mortality by seven days to align with weekly reporting cycles, account for delays in death reporting, and approximate the time individuals take to perceive and react to evolving risk information through media and social networks. To account for the persistence of mobility patterns, we included lagged mobility (*Mobility_i,t-7_*, *Mobility_i,t-14_*), which control for short-term mobility inertia and ensure that our estimates capture dynamic adjustments to risk perception rather than residual behavioral trends. We further control for county-fixed effects (*α_i_*) to account for time-invariant unobserved heterogeneity across counties. We further control for month-fixed effects (*δ_m_*) and day-of-the-week fixed effects (*λ_d_*) to remove systematic and broader seasonal trends in mobility that affect all counties simultaneously. However, these global time effects cannot account for heterogenous local trajectories in mobility driven by county-specific factors (i.e., economic activity, policy enforcement, or behavioral adaptation). To further isolate short-run behavioral responses from gradual, county-specific trends, we include a county-specific linear time trend (*γ_i_t*), which allows, each county’s baseline mobility to evolve independently over time. This specification distinguishes short-run behavioral responses to changes in local mortality -the identifying variation-from slower-moving local shifts in mobility driven by structural factors. Additionally, we included policy variables - *Shelter in Place_it_* and *Mask* − *Mandate_it_* - to isolate the effect of risk perception (proxied by mortality) on private preventive behaviors from reductions in mobility that were directly mandated by government interventions. By controlling for these policies, we ensure that our estimates capture voluntary behavioral responses to risk perception rather than government-imposed restrictions on mobility. The coefficient *β*_1_ captures the semi-elasticity of voluntary behavioral responsiveness to mortality-driven risk perception, net of both persistent regional differences and gradual temporal trends. Identification arises from short-run, within-county fluctuations in mortality and mobility around each county’s evolving baseline rather than from broader behavioral dynamics or gradual shifts in collective behavior. Standard errors are clustered at the county level to account for serial correlation in unobserved shocks over time. (See Appendix for robustness check).

#### Heterogeneity, Temporal Dynamics, and Policy Effects

Mobility across Retail & Recreation, Grocery & Pharmacy, Workplaces, Transit Stations, Parks, and Residential may reflect different types of private preventive behaviors. To capture these distinctions, we separately examined them, each serving as an individual outcome variable in separate regressions. Retail & Recreation mobility reflects non-essential activities and captures individuals’ willingness to visit leisure-related locations despite potential exposure risks. Grocery & Pharmacy mobility represents essential movement, where individuals must balance necessary outings with risk avoidance. Transit Station mobility involves movement with limited capacity for social distancing, yet it remains an essential form of transportation for many. Park mobility differs from the others, as outdoor spaces were widely perceived as safer environments, often serving as a behavioral substitution for riskier indoor activities rather than direct risk avoidance. Residential mobility, measured as time spent at home, is a direct proxy for the extent individuals engage in complete risk avoidance by minimizing exposure to public spaces. All mobility measures are log-transformed and normalized by county population to account for differences in scale across counties. Next, we investigate whether the sensitivity of mobility behavior to risk perception weakens as the pandemic progresses by interacting with time our key independent variable, lagged deaths per 10,000 residents. This effect reflects pandemic fatigue.

Finally, we examined how public health policies mediate behavioral responses to risk perception by introducing interaction terms between lagged deaths and policy variables. We first interacted lagged deaths with shelter-in-place orders and then in a separate regression with mask mandates and finally include a triple interaction between both policies to analyze their combined effect. These interactions allow us to determine whether public policies amplify, dampen, or have no effect on voluntary behavioral responses to risk perception. If the estimated interaction terms are negative, it would suggest that the presence of formal restrictions reinforces mobility reductions beyond what is driven by risk perception alone. Conversely, if the interaction effects are positive or insignificant, it would indicate that individuals primarily rely on their assessment of risk rather than government-imposed policies when adjusting mobility behaviors.

In all these additional regressions, we maintained the same set of controls, county and time-fixed effects, and clustered standard errors as in our main specification.

## Results

Our estimates capture short-run, within-county behavioral adjustments to perceived mortality risk, reflecting voluntary changes in mobility rather than long-term structural or policy-driven trends. Across 1,206 counties and 1.16 million county-day observations, a one-unit increase in COVID-19 deaths per 10,000 residents (lagged seven days) is associated with a 4.8% reduction in total mobility (p < 0.001), reflecting collective adjustments to perceived risk. ^1^ However, this average effect conceals distinct behavioral regimes across pandemic stages.

During the early pandemic, the estimated coefficient on mortality-driven risk perception is –0.032 (p < 0.01), corresponding to a 3.1% decline in mobility per additional death per 10,000 residents (7-day lag). This attenuated behavioral response contrasts sharply with the stronger policy effect, as shelter-in-place mandates during this stage are associated with a 10.7% reduction in mobility—suggesting that early behavioral changes were largely policy- and fear-driven, rather than tied to observed local mortality. In the risk-compensation period, by contrast, the behavioral elasticity strengthened considerably (β = –0.081, p < 0.001), equivalent to a 7.8% reduction in mobility per additional death per 10,000 residents, even as the effect of shelter-in-place orders weakened to –2.0%. This inversion supports our central hypothesis: as uncertainty declined and individuals gained experience and information about the virus, behavioral responses became more calibrated and mortality-sensitive, reflecting adaptive private choices rather than broad panic or mandated compliance. By the post-vaccination period, the mortality–mobility association was statistically insignificant (β = 0.007, p = 0.24), consistent with behavioral desensitization as perceived risk diminished amid widespread vaccine coverage (Table 1 and Figure 1).

**Figure 1.**
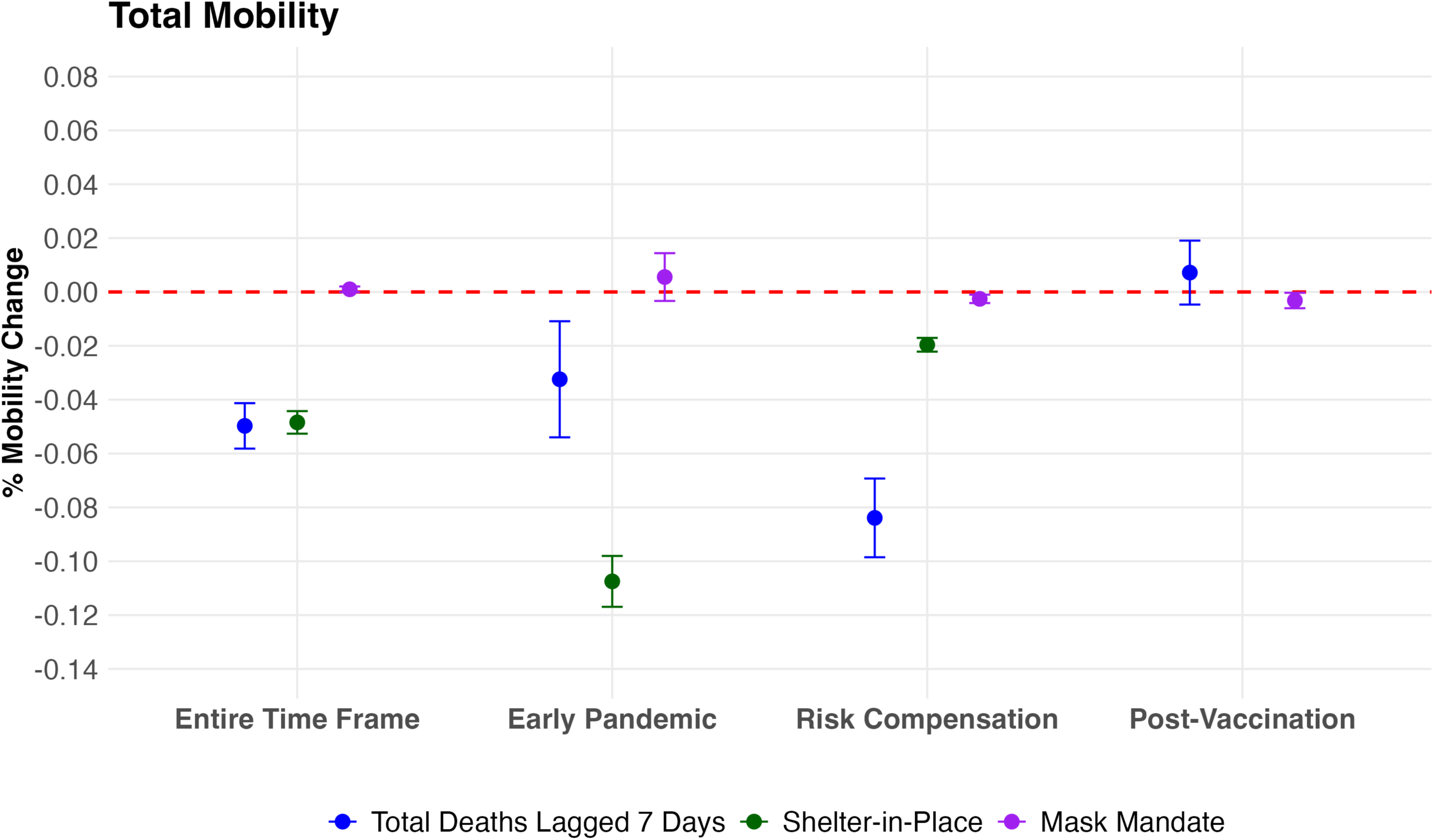
Estimated Effects of Mortality driven Risk Perception and Public Health Policies on Total Mobility Across Pandemic Phases.

**Table 1.** Effect of Mortality driven Risk Perception and Public Health Policies on Total Mobility Across Pandemic Phases. Main regression results across defined stages of the pandemic. Outcome: total mobility.

|  | Entire Time Frame | Early Pandemic | Risk Compensation | Post-Vaccination |
| --- | --- | --- | --- | --- |
| **Deaths per 10,000 (7-day lag)** | −0.050<br>(CI: −0.058, −0.041)<br>p < 0.001 | −0.032<br>(CI: −0.054, −0.011)<br>p = 0.003 | −0.084<br>(CI: −0.099, −0.069)<br>p < 0.001 | 0.007<br>(CI: −0.005, 0.019)<br>p = 0.235 |
| **Shelter-in-Place Active** | −0.048<br>(CI: −0.053, −0.044)<br>p < 0.001 | −0.107<br>(CI: −0.117, −0.098)<br>p < 0.001 | −0.020<br>(CI: −0.022, −0.017)<br>p < 0.001 | — |
| **Mask Mandate Active** | 0.001<br>(CI: −0.000, 0.002)<br>p = 0.065 | 0.006<br>(CI: −0.003, 0.014)<br>p = 0.221 | −0.003<br>(CI: −0.004, −0.001)<br>p = 0.001 | −0.003<br>(CI: −0.006, −0.000)<br>p = 0.030 |
| **County FE** | ✓ | ✓ | ✓ | ✓ |
| **Time FE (Month, Day-of-week)** | ✓ | ✓ | ✓ | ✓ |
| **Lagged Mobility Controls** | ✓ | ✓ | ✓ | ✓ |
| **County-specific time trends** | ✓ | ✓ | ✓ | ✓ |

To further unpack these dynamics, we examine how observed mortality risk influenced different types of mobility during the risk-compensation period. Because each form of mobility represents a distinct private preventive behavior, these disaggregated outcomes provide deeper insight into how communities adjusted their collective routines in response to perceived risk. We found that higher mortality risk led to significant reductions in Retail & Recreation (–5.15%, p < 0.001) and Transit Station (–5.33%, p < 0.001) mobility—settings typically associated with discretionary and high-contact activities. These categories show the strongest behavioral adjustments, likely reflecting efforts to limit exposure in non-essential and crowded environments. Workplace mobility also declined substantially (–7.53%, p < 0.001), suggesting that even work-related activities were scaled back as risk increased, possibly due to expanded remote work or voluntary avoidance of shared indoor spaces. Grocery & Pharmacy mobility was largely unaffected by local mortality risk (–0.41%, p = 0.21), reflecting the essential nature of these trips and limited flexibility to substitute them. Parks mobility, however, remained unchanged (–1.7%, p = 0.32). This null effect is both intuitive and meaningful: outdoor spaces were broadly perceived as lower-risk environments during the pandemic. Finally, residential mobility increased by 1.5% (p < 0.001), reflecting a shift toward time spent at home—a behavioral response consistent with private risk avoidance. Together, these findings reveal that communities selectively adjusted their movement patterns based on perceived exposure risk, reducing non-essential mobility while preserving essential and low-risk activities (Table 2 and Figure 2). Full category-specific results across all pandemic phases are provided in Appendix Section 4, which details how mortality-driven risk perception and public health policies differentially influenced discretionary, essential, and residential forms of mobility.

**Figure 2.**
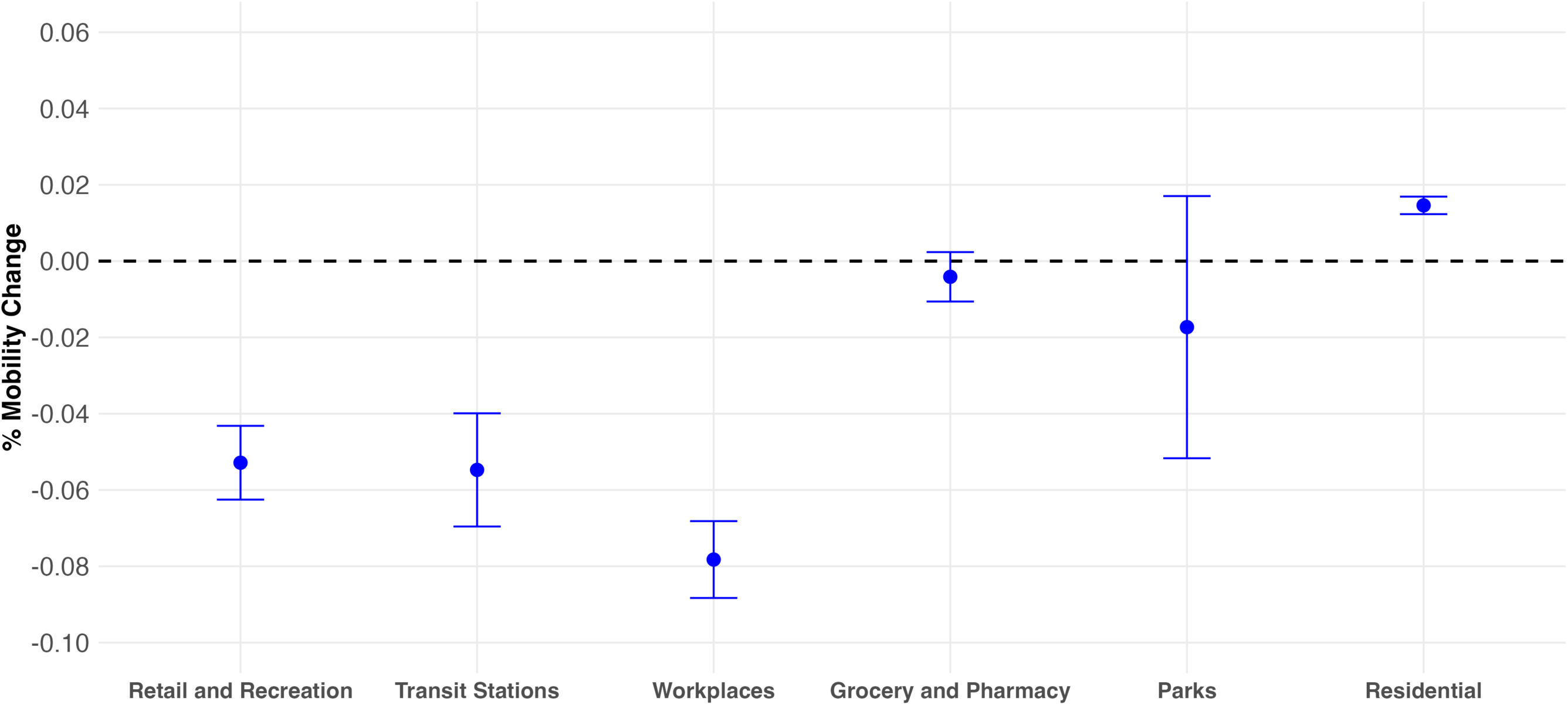
Category-Specific Behavioral Responses to Mortality driven Risk Perception During the Risk Compensation Period.

**Table 2.** Category-Specific Behavioral Responses to Risk Perception During the Risk Compensation Period. Regression results for each mobility category as outcome. Period: Risk Compensation.

|  | Retail & Recreation | Transit Stations | Workplaces | Grocery & Pharmacy | Residential | Parks |
| --- | --- | --- | --- | --- | --- | --- |
| **Deaths per 10,000 (7-day lag)** | -0.053<br>(CI: -0.063, -0.043)<br>p < 0.00 | -0.055<br>(CI: -0.070, -0.040)<br>p < 0.001 | -0.078<br>(CI: -0.088, -0.068)<br>p < 0.001 | -0.004<br>(CI: -0.011, 0.002)<br>p = 0.213 | 0.015<br>(CI: 0.012, 0.017)<br>p < 0.001 | -0.017<br>(CI: -0.052, 0.017)<br>p = 0.323 |
| **Shelter-in-Place Active** | -0.023<br>(CI: -0.026, -0.020)<br>p < 0.001 | -0.009<br>(CI: -0.013, -0.004)<br>p < 0.001 | -0.027<br>(CI: -0.030, -0.025)<br>p < 0.001 | -0.003<br>(CI: -0.005, -0.000)<br>p = 0.023 | 0.003<br>(CI: 0.003, 0.003)<br>p < 0.001 | -0.011<br>(CI: -0.019, -0.003)<br>p = 0.006 |
| **Mask Mandate Active** | -0.012<br>(CI: -0.014, -0.010)<br>p < 0.001 | -0.005<br>(CI: -0.008, -0.001)<br>p = 0.008 | 0.002<br>(CI: -0.000, 0.004)<br>p = 0.121 | -0.008<br>(CI: -0.009, -0.006)<br>p < 0.001 | 0.001<br>(CI: 0.001, 0.002)<br>p < 0.001 | -0.015<br>(CI: -0.023, -0.007)<br>p < 0.001 |
| **County FE** | ✓ | ✓ | ✓ | ✓ | ✓ | ✓ |
| **Time FE (Month, Day-of-week)** | ✓ | ✓ | ✓ | ✓ | ✓ | ✓ |
| **Lagged Mobility Controls** | ✓ | ✓ | ✓ | ✓ | ✓ | ✓ |
| **County-specific time trends** | ✓ | ✓ | ✓ | ✓ | ✓ | ✓ |

Finally, we examine whether behavioral responses to observed mortality risk varied over time and across policy environments. In the first specification, we found that the effect of observed mortality risk on mobility weakens over time, with the interaction term indicating a 0.5 percentage point attenuation per additional month (p < 0.001) (Table 3). This result is consistent with a process of pandemic fatigue [24], [25], [26].

**Table 3.**
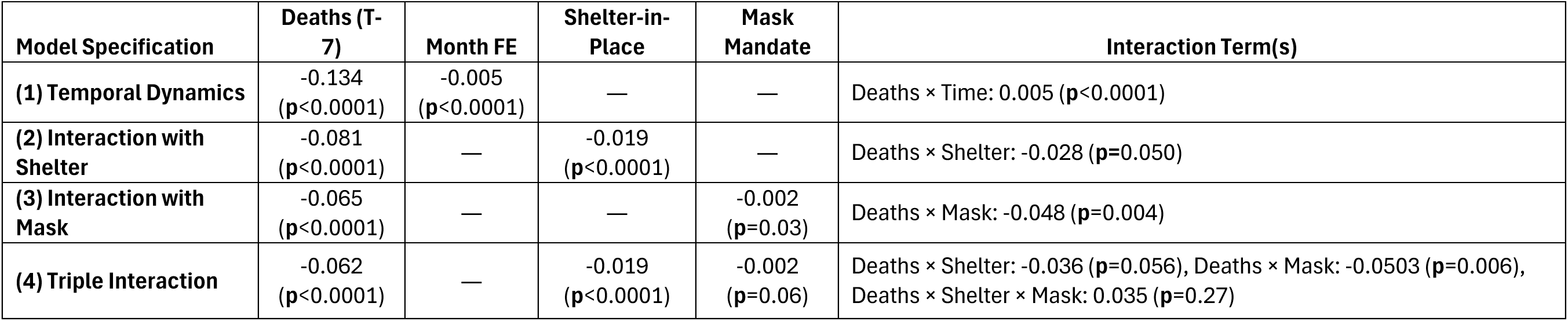
Interaction Effects: Temporal Dynamics and Policy Moderation of Risk Responses. Models testing whether behavioral responses to deaths vary by time and policy environment. Period: Risk Compensation.

Turning to policy moderation, we observed that community behavioral response becomes stronger when shelter-in-place orders are active. Specifically, the interaction term suggests an additional 2.8% reduction in mobility in response to observed mortality risk during periods with active shelter-in-place policies (p = 0.050). Similarly, mask mandates amplify the effect of observed mortality risk, with an additional 4.7% reduction in mobility (p = 0.004). These findings suggest that formal policy interventions may reinforce individual risk perception, prompting a stronger private preventive action. The triple interaction between mortality risk, shelter-in-place, and mask mandates was small (3.5%) and statistically insignificant (p = 0.27), indicating no additional synergistic effect (Table 3).

## Discussion

Our contribution makes clear that preventative behavior is not passive—it is endogenously reactive to perceived health risk and evolves as the informational and epidemiological landscape shifts. We show that communities don’t simply increase the demand for private preventive behaviors in the abstract; rather, they recalibrate specific behaviors based on perceived risk exposure and necessity. Importantly, our disaggregated analysis across different types of mobility domains—ranging from discretionary (e.g., Retail & Recreation) to essential (e.g., Grocery) to substitutive (e.g., Parks)—reveals how different categories of preventive action respond to risk in distinct ways. This heterogeneity uncovers the behavioral logic behind risk-avoidant actions, showing that communities selectively recalibrated collective routines in response to evolving local mortality signals.

Equally important, this study shows that public health policies do not merely impose restrictions but also shape private behavior by modulating the strength of voluntary responses. Rather than crowding out risk-avoidant behavior, mandates such as shelter-in-place orders and mask requirements amplify it—suggesting that these policies serve not just as constraints but as catalysts for heightened vigilance. These findings demonstrate that top-down interventions reinforce, rather than replace, private protective behaviors—establishing a mutually reinforcing relationship between institutional mandates and individual action. While prior studies have focused on estimating the average effects of mandates on behavior or mortality (e.g., [18], [19], [27], [28], [29]), our analysis reveals a more dynamic interaction as it suggests that mandates not only act as external restrictions that limit movement or prescribe actions but as contextual amplifiers of perceived risk. While Goolsbee and Syverson (2021) highlight the importance of voluntary behavior and Chernozhukov et al. (2021) decompose average policy effects through behavioral channels, our analysis goes further by modeling how public health mandates amplify individuals’ responsiveness to perceived mortality risk. Unlike prior studies limited to the early months, we examine behavioral dynamics across multiple pandemic stages to capture how responsiveness evolved with shifting informational and epidemiological contexts [30].

In parallel, instead of studying risk compensation behaviors that occur after protection is adopted, we examine how individuals proactively adjust preventive behaviors in response to real-time, objective risk signals. This provides a distinct perspective on behavioral adaptation, in contrast to most of the existing literature in risk compensation behavior, which focuses on reduced vigilance or risk-taking. For instance, behavioral offsets following PrEP initiation or HIV treatment (e.g., [11], [12], [13], [14]), or after vaccination or mask mandates during COVID-19 (e.g., [15], [16], [17], [31], [32]). Rather than assuming behavior changes only once protection is secured, we show that individuals dynamically respond to fluctuations in perceived threat—adjusting their routines in anticipation of risk, not just in reaction to intervention ([19], [33]).

Finally, we introduce a novel methodological framework that integrates SafeGraph’s absolute mobility data with Google’s category-specific trends to reconstruct daily, domain-specific levels of private movement. Prior studies using mobility data typically rely on relative changes to pre-pandemic baselines (Google Mobility data) or absolute-level data without information on mobility across different categories (e.g., [17], [18], [19], [34], [35]). In contrast, our fusion of data sources allows us to recover the absolute level of mobility across six distinct behavioral domains—enabling a more behaviorally grounded analysis of how different types of private preventive actions evolve in response to perceived risk. Using this enriched dataset, we also empirically estimate pandemic fatigue as a declining responsiveness of mobility to mortality-driven risk perception. While the notion of behavioral fatigue has been theorized and estimated (e.g., [36], [37], [38], [39]), our analysis directly estimates its effect: responsiveness weakened by ∼1 percentage point per month, underscoring the erosion of vigilance over time and the need for adaptive policy strategies.

Several limitations warrant acknowledgment. First, unmeasured factors—such as local media, political attitudes, or institutional trust—may shape both behavior and its responsiveness to risk, as perceived risk reflects subjective interpretation and how populations internalize mortality signals. Second, while mobility is a useful proxy for preventive action, it excludes behaviors like masking or distancing within settings. Third, county-level analysis captures aggregate patterns but not individual heterogeneity or motivations. Despite these limitations, our findings provide macro-level evidence on the dynamic, mutually reinforcing interaction between private behavior and public policy.

### Public Health Implications

This study highlights that community behavior is not passive but dynamically adjusts to evolving perceptions of risk. We find that mortality-driven risk perception shaped strong preventive responses that weakened over time, while public health mandates amplified rather than displaced these actions. Policies do not operate in isolation—they interacted with community responses to mortality-driven risk perception, reinforcing vigilance during the pandemic but losing strength as fatigue eroded collective responsiveness. Recognizing this interplay is critical for designing interventions that remain effective beyond their immediate, direct effects.

Our framework provides a practical tool for anticipating how shifts in perceived protection— whether from new treatments, preventive technologies, or policy announcements—can alter private behavior in ways that reshape population-level outcomes.

This study underscores the necessity of integrating behavioral adaptation into policy design. As we prepare for future health emergencies, this framework can inform more responsive, human-centered interventions that align with how people actually perceive and act upon risk in real time. The behavioral plasticity revealed here is not a liability but a resource to be understood and leveraged.

## Data Availability

All data is publicly available

## Data and Code Availability

All data used in this study are publicly available. The code used for the calibration of mobility data is available upon request, and all other analysis code is available upon request from the corresponding author.

## Ethics Approval

Not applicable.

## Appendix

### 1. Mobility Data

#### Data Sources

We utilized two complementary data sources to measure mobility behavior comprehensively: Google’s COVID-19 Community Mobility Reports and SafeGraph Mobility Data.

*Google’s dataset* provides daily, anonymized percentage changes in visits across key categories, including Retail & Recreation, Grocery & Pharmacy, Workplaces, Transit Stations, Parks, and Residential (time spent at home) for the entire study timeframe. These mobility changes are reported relative to a pre-pandemic baseline, which is defined as the median value for each weekday (Monday through Sunday) from January 3 to February 6, 2020 [40]. This weekday-specific baseline accounts for systematic differences in movement patterns across days of the week, ensuring that mobility trends are contextualized over time.

*SafeGraph* tracks mobility using origin-to-destination visits based on anonymized device location data. A visit is recorded when a device remains at both the origin and the destination for at least one minute, ensuring that movement between locations reflects actual visits rather than transient signals [41]. This approach allows for the identification of individual-level mobility trends while preserving privacy. By leveraging aggregated location signals, these mobility data provide insights into patterns of movement and how they change over time in response to external factors such as public health policies and perceived risk.

SafeGraph’s dataset, available until March 2021, provides absolute mobility levels at the county level, capturing origin-to-destination visitor flows within counties adjusted for population distribution. Unlike Google’s dataset, which reports percentage changes by location category, SafeGraph offers absolute mobility figures without categorical breakdowns.

Each dataset presents specific limitations. Google’s mobility data measures percentage changes relative to a pre-pandemic baseline rather than absolute movement levels. SafeGraph, while providing absolute mobility levels, is restricted to data availability only until March 2021 and lacks category-specific mobility breakdowns. The absence of categorical detail in SafeGraph data is particularly relevant, as different types of mobility reflect distinct private preventive behaviors. Given these constraints, we integrate both datasets to construct a more robust and consistent measure of mobility, leveraging SafeGraph’s absolute mobility levels for baseline estimates and Google’s category-specific percentage changes for mobility trends across the whole study period.

#### Integration of Data Sources: Mobility Measurement

To generate a consistent, category-specific measure of absolute mobility, we integrated SafeGraph’s total absolute mobility levels with Google’s category-specific mobility trends, which report *relative changes* in mobility compared to a pre-pandemic baseline. First, we estimated daily baseline absolute mobility for each county using SafeGraph data from the pre-pandemic period (January 3–February 6, 2020). We then calibrated the fraction of this total baseline mobility allocated to each domain (e.g., Workplaces, Retail & Recreation) using Google’s relative mobility trends and SafeGraph data. Because mobility patterns differ between weekdays and weekends, we estimated separate calibrated shares for each: for weekdays, 10% each to Retail & Recreation, Transit, and Grocery & Pharmacy, 64.85% to Workplaces, and 5.06% to Parks; for weekends, shares shift to 15.29% for Retail & Recreation, 15.13% for Grocery & Pharmacy, 17.69% for Transit, 46.82% for Workplaces, and 5.07% for Parks. Finally, we applied Google’s daily percentage changes to these calibrated baselines to reconstruct daily absolute mobility by category across the study period. This method ensures internal consistency and captures meaningful variation across both time and activity type, while enabling us to recover absolute, daily mobility levels by category—a critical feature for analyzing the intensity and timing of private preventive behaviors across distinct domains, each representing different forms of risk-mitigating action. A detailed explanation of the calibration procedure and validation is provided in the next section.

#### Calibrating Category-Specific Mobility at Baseline

To estimate baseline category-specific mobility, we calibrate the fraction of total mobility attributed to Retail & Recreation, Transit, Grocery & Pharmacy, Workplaces, and Parks in the baseline period. This calibration addresses the limitation of SafeGraph data, which provides only total mobility without categorical breakdowns. By leveraging Google’s mobility percentage changes and SafeGraph’s county-level total mobility data, we derive the fraction of total mobility that corresponds to each category in the baseline period, ensuring consistency between both datasets.

#### Timeframe for Calibration Model

The calibration procedure is conducted using mobility data from February 15 to March 15, 2020, a period preceding widespread lockdown measures. This timeframe is chosen to ensure that the model captures pre-lockdown behavioral shifts, minimizing disruptions in mobility patterns caused by the onset of the COVID-19 pandemic and the subsequent implementation of public health policies.

#### Optimization Setup & Methodology

The calibration process estimates category-specific mobility shares by minimizing the squared error between observed log-mobility (SafeGraph total mobility), which serves as the target variable, and the reconstructed log-mobility, derived from category-weighted SafeGraph baselines. This is achieved using the L-BFGS-B algorithm, a constrained optimization approach that ensures the estimated category probabilities remain non-negative and sum to a value no greater than one.

Let:

- *c* index counties
- *t* index time (days)
- *d*(*t*) denote whether day *t* is a weekday or weekend.
- *k* ∈ {*retail&recreation, transit stations, workplaces, grocery&pharmacy, parks*}
- 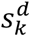 denote the category share for category *k* for day type *d*, which we calibrate.
- 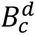 denote the baseline total mobility per 10,000 people in county *c* on day type *d*, derived from SafeGraph.
- Δ*_k_*_,*ct*_ denote the Google-reported percentage change from baseline for category *k* in county *c* at time *t*.
- 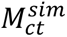 denote the Simulated Total Mobility per 10,000 people in county *c* at time *t*.

Then, the simulated mobility is defined as:

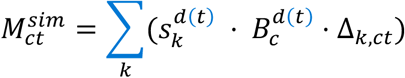

By combining these elements, we reconstruct total mobility for each time *t* included in the calibration period (February 15 to March 15, 2020). Note that we calibrate a distinct share for each of the five mobility categories—Retail & Recreation, Transit Stations, Workplaces, Grocery & Pharmacy, and Parks—separately for weekdays and weekends. Since category shares must sum to one within each day type, we fix the Parks share and calibrate the remaining four, yielding a total of 8 parameters (4 categories × 2 day types). This weekday/weekend-specific structure is essential because Google’s mobility percentage changes are defined relative to day-specific baseline values, capturing systematic pre-pandemic movement patterns. Calibrating shares separately for weekdays and weekends ensures that reconstructed mobility aligns with the structure of the Google mobility data while preserving the internal consistency of total mobility estimates.

#### Stochastic Initialization

To mitigate convergence issues and avoid local minima in the optimization process, we employ a stochastic initialization strategy. Specifically, we randomly sample 5,000 sets of initial values for category probabilities. The best-fitting set is selected based on the lowest calibration error, ensuring an optimal allocation of total mobility across categories.

#### Final Calibrated Output

The final calibrated category shares reflect distinct weekday versus weekend patterns. For weekdays (Monday through Friday), 10% of total mobility is attributed to each of Retail & Recreation, Transit, and Grocery & Pharmacy, 64.85% to Workplaces, and the remaining 5.06% to Parks. For weekends (Saturday and Sunday), we observe increased mobility shares for Retail & Recreation (15.29%), Grocery & Pharmacy (15.13%) and Transit (17.69%), with a corresponding reduction in Workplaces (46.82%), while Parks maintain a fixed residual share of approximately 5.07%. These estimates are reported in the table below:

|  | Retail & Recreation | Transit Stations | Workplaces | Grocery & Pharmacies | Parks |
| --- | --- | --- | --- | --- | --- |
| Weekday | 10% | 10% | 64.85% | 10% | 5.06% |
| Weekend | 15.29% | 17.69% | 46.82% | 15.13% | 5.07% |

To validate the fit of our calibration model, we use the estimated category shares presented above to reconstruct simulated total mobility, which we then compare against the observed SafeGraph total mobility for the calibration period. As shown in Figure A1, the simulated mobility closely tracks the observed national average, capturing key trends and fluctuations.

**Figure A1:**
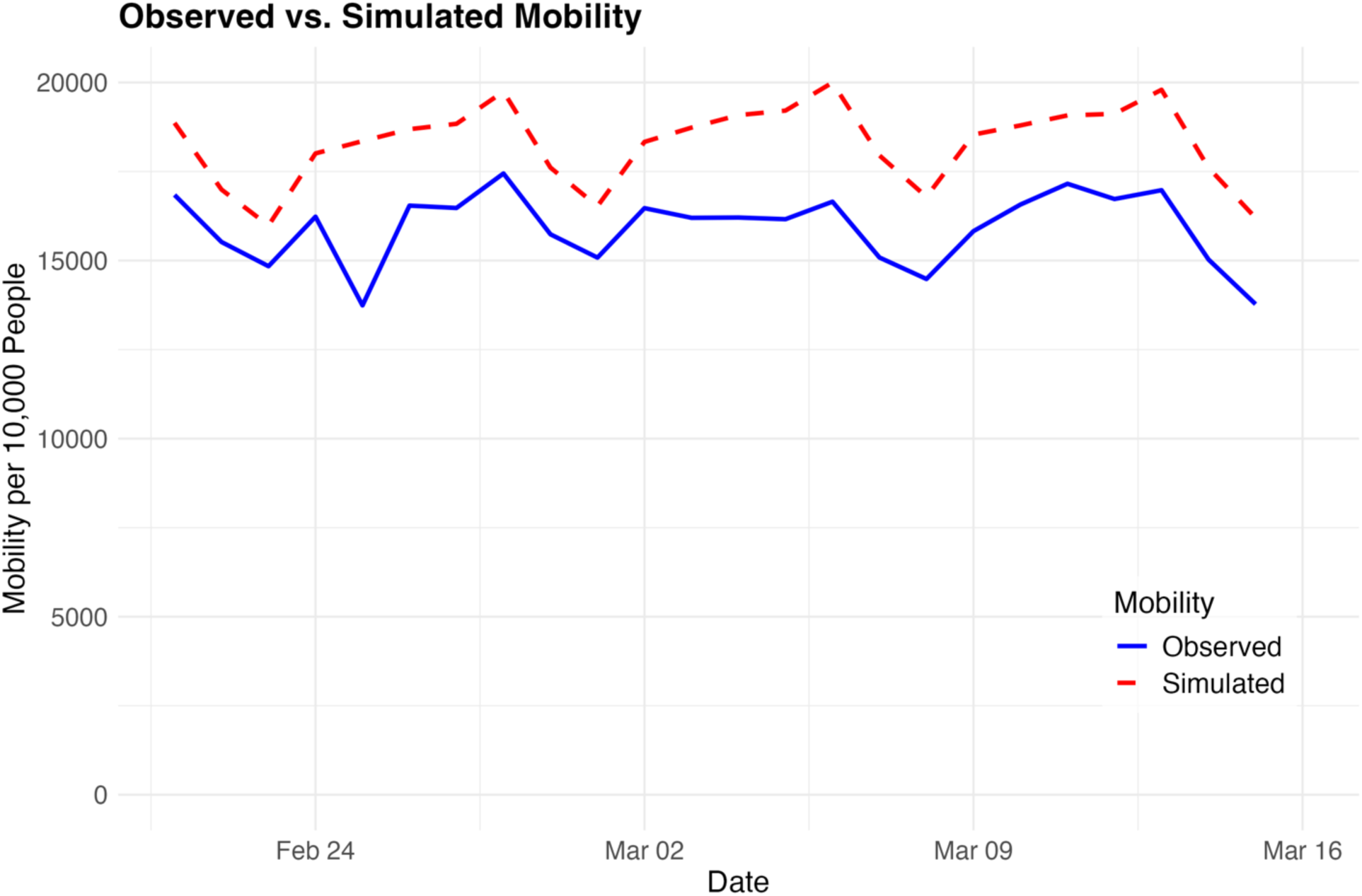
Observed vs. Simulated Total Mobility.

#### Out-of-Sample Validation Exercise

To assess the validity of our calibration approach, we conduct an out-of-sample validation exercise. Since SafeGraph provides observed total mobility only until March 2021, we leverage the calibrated category-specific shares to reconstruct daily total mobility beyond this period. Specifically, we use the estimated shares to generate category-specific mobility for the out-of-sample period, which we then aggregate to obtain total mobility estimates. These reconstructed mobility trajectories are compared against the observed total mobility reported by SafeGraph.

As shown in Figure A2, the trends, relative changes, and overall patterns of our reconstructed total mobility align closely with the observed SafeGraph mobility data. This alignment indicates that our calibration approach effectively captures the underlying mobility dynamics, reinforcing the robustness of our methodology. The consistency between the estimated and observed mobility suggests that our calibrated shares provide a reliable approximation of category-specific movement patterns, even beyond the calibration period.

**Figure A2:**
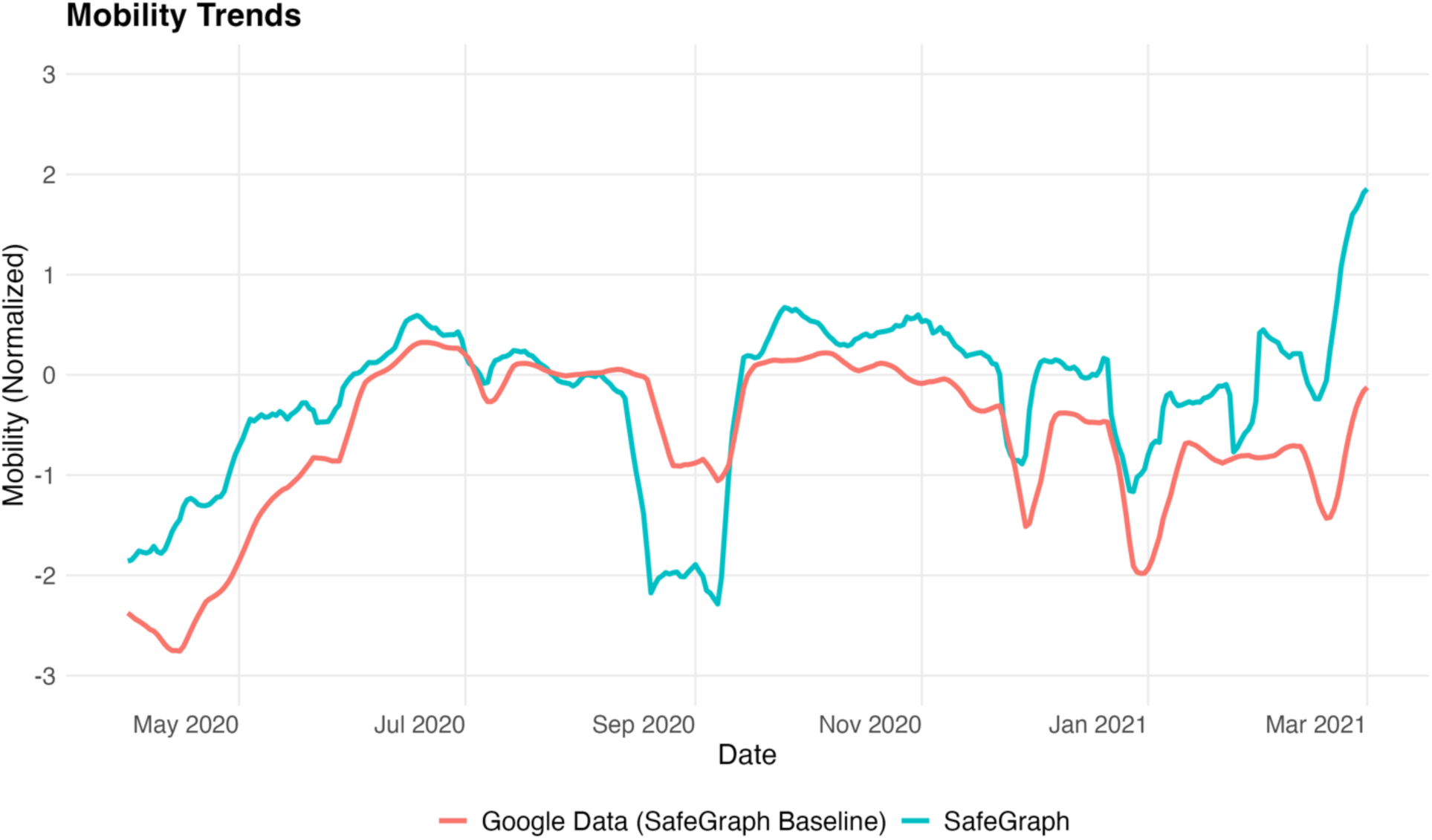
External validation: Mobility Trends.

### 2. Pandemic Period Classification

Given the evolving nature of the pandemic, we divided our analysis into four stages, reflecting changes in public knowledge about the virus, levels of uncertainty, perceived risk, and patterns of individual behavioral response—defined by contextual shifts rather than fixed epidemiological thresholds.

1.- Early Pandemic (February 15 – April 30, 2020): This phase represents the initial period of widespread uncertainty and crisis-driven behavioral responses. The first confirmed COVID-19 case in the United States was reported in Washington State on January 20, 2020. The World Health Organization (WHO) declared a Public Health Emergency of International Concern on January 30, 2020, and officially classified COVID-19 as a pandemic on March 11, 2020. The United States declared a national emergency on March 13, 2020. During this phase, public uncertainty was at its peak, with widespread fear driven by overwhelmed hospitals and a lack of information on the virus’s transmission and severity. By the end of April, at the peak, approximately 88.3% of U.S. counties had active shelter-in-place orders, and restrictions were widespread. Mortality rates were high, with an estimated case fatality ratio of ∼0.05. Preventive behaviors were driven by general uncertainty rather than calculated risk assessment, as individuals reduced mobility across all domains regardless of risk perception (mortality trends).

2.- Risk Compensation Period (May 1, 2020 – February 28, 2021): As public knowledge of COVID-19 increased, risk perception shifted from generalized fear to a more structured assessment based on real-time mortality trends. During this phase, individuals adjusted their private preventive behaviors in response to evolving risk signals. Policy restrictions also changed, with shelter-in-place orders declining rapidly in early May, leaving only ∼12% of U.S. counties with active orders by June 2020. Individuals’ reliance on mortality data to guide risk perception grew as they became more informed [20], [21], [22]. While mobility gradually increased, it remained approximately 20% below pre-pandemic levels, and the case fatality ratio dropped to ∼0.01.

3.- Vaccination Rollout / Transitional Period (March 1 – May 31, 2021): By March and April 2021, vaccines became available to all adults in most states, but uptake varied, leading to heterogeneity in behavioral responses. This period represents a transitional phase where behavioral adjustments were less clearly defined, as individuals responded to both increasing vaccine availability and evolving risk perceptions.

4.- Post-Vaccination Phase (June 1 – November 30, 2021): By June 1, 2021, nearly half of the U.S. adult population had received at least one vaccine dose, and coverage continued to expand. Mortality rates remained low during this period, and mobility trends neared pre-pandemic levels. With declining perceived risk, individuals may have reassessed the need for precautionary measures. No counties had active shelter-in-place orders by this stage, and the case fatality ratio had fallen below 0.01.

We conducted our empirical analysis separately for each of these stages to examine how responsiveness to mortality risk evolved as the pandemic progressed. This structure enabled us to isolate shifts in preventive behavior under varying informational and epidemiological conditions.

### 3. Robustness Checks

As robustness checks, we implemented three alternative specifications to assess potential model dependence in our main analysis of total mobility during the risk compensation period:

1. We added a 21-day lag of mobility to account for longer-run behavioral persistence;
2. We included a 14-day lag of mortality to account for temporal correlation in deaths and test for sensitivity to longer risk signal windows; and
3. We removed lagged mobility to test whether the estimated effects were driven by serial correlation rather than responses to mortality signals.

The results remain robust across all alternative specifications. The main estimated effect of observed mortality on total mobility was –8.0 % (p < 0.001). Including a 21-day lag of mobility yields a slightly attenuated effect of –6.8 % (p < 0.001), while adding a 14-day mortality lag produces a coefficient equivalent to a –5.5 % change (p < 0.001), consistent with temporally correlated risk perception. Removing lagged mobility amplifies the estimated effect to –12.1 % (p < 0.001), suggesting that the baseline specification is conservative and not driven by behavioral persistence.

### 4. Category-Specific Behavioral Responses Across Pandemic Phases

This appendix presents category-specific estimates of how mortality-driven risk perception and public health policies shaped mobility behaviors across different stages of the COVID-19 pandemic. Each figure reports coefficients and 95% confidence intervals from the main log-linear specification, controlling for county and time fixed effects (month and day-of-week), lagged mobility, and county-specific linear time trends. The figures display how the effects of mortality-driven risk perception and public health interventions evolved across activity domains—reflecting discretionary, essential, and residential forms of private preventive behavior.

**Figure A3.**
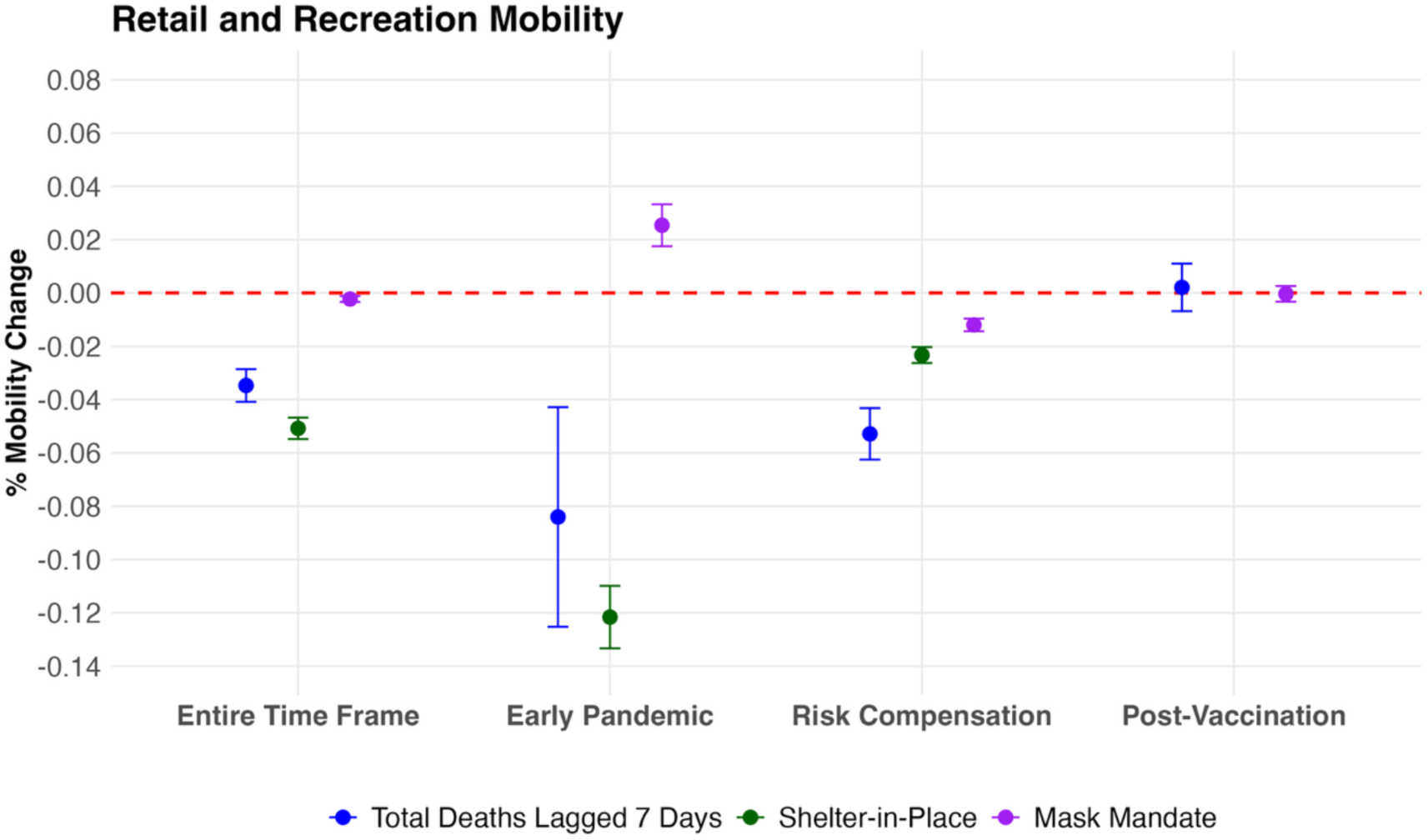
Estimated Effects of Mortality-Driven Risk Perception and Public Health Policies on **Retail and Recreation Mobility** Across Pandemic Phases

**Figure A4.**
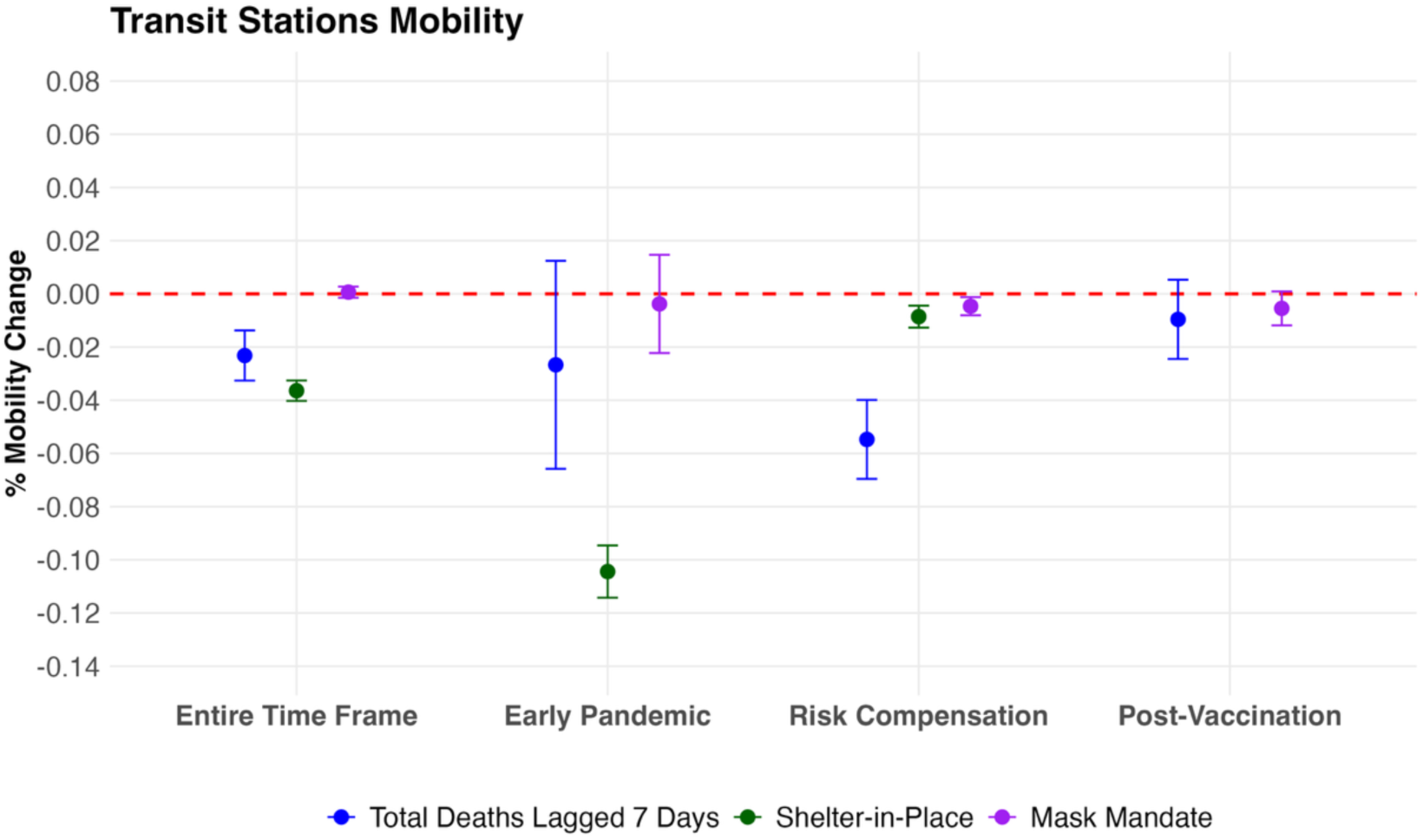
Estimated Effects of Mortality-Driven Risk Perception and Public Health Policies on **Transit Station** Mobility Across Pandemic Phases

**Figure A5.**
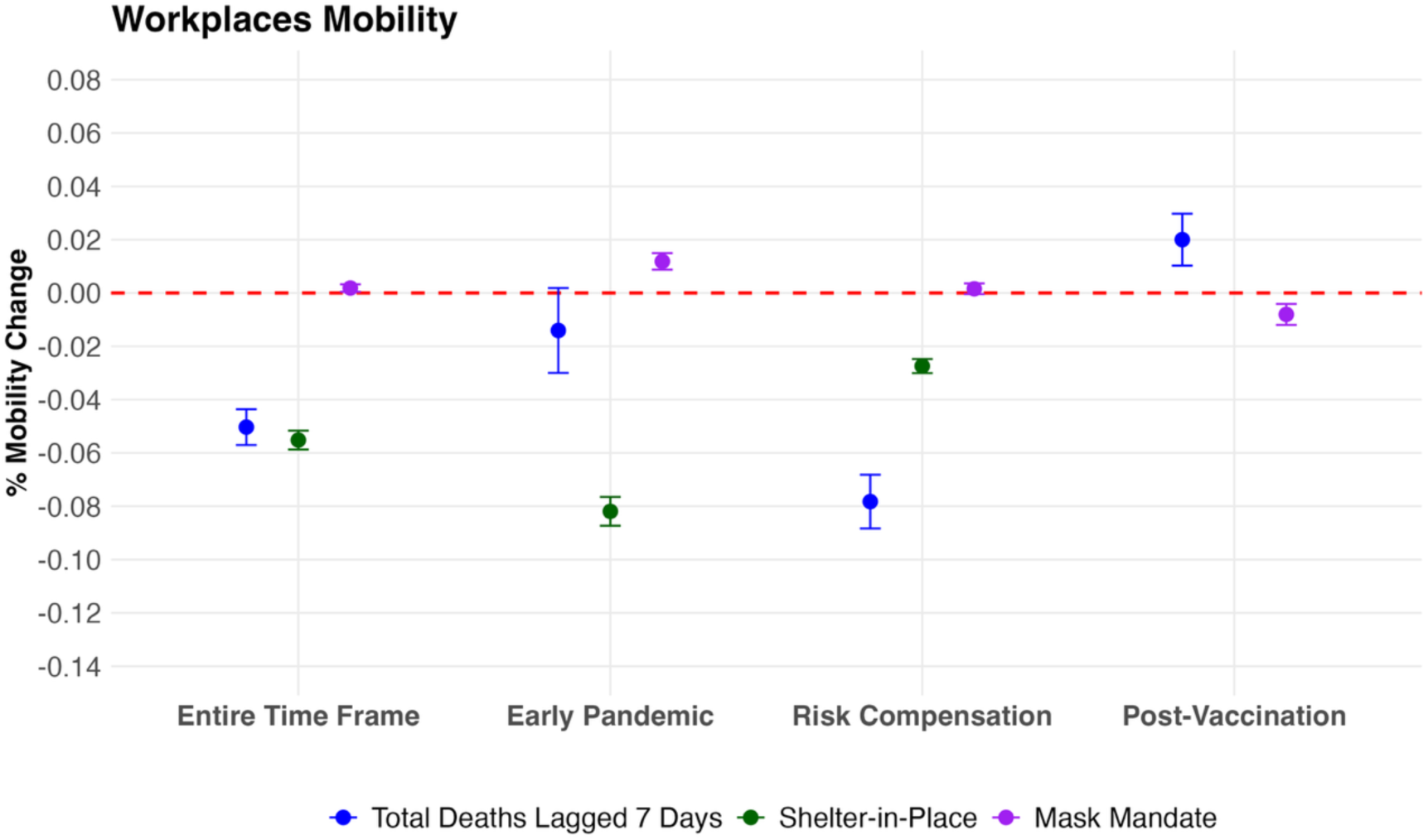
Estimated Effects of Mortality-Driven Risk Perception and Public Health Policies on **Workplace** Mobility Across Pandemic Phases

**Figure A6.**
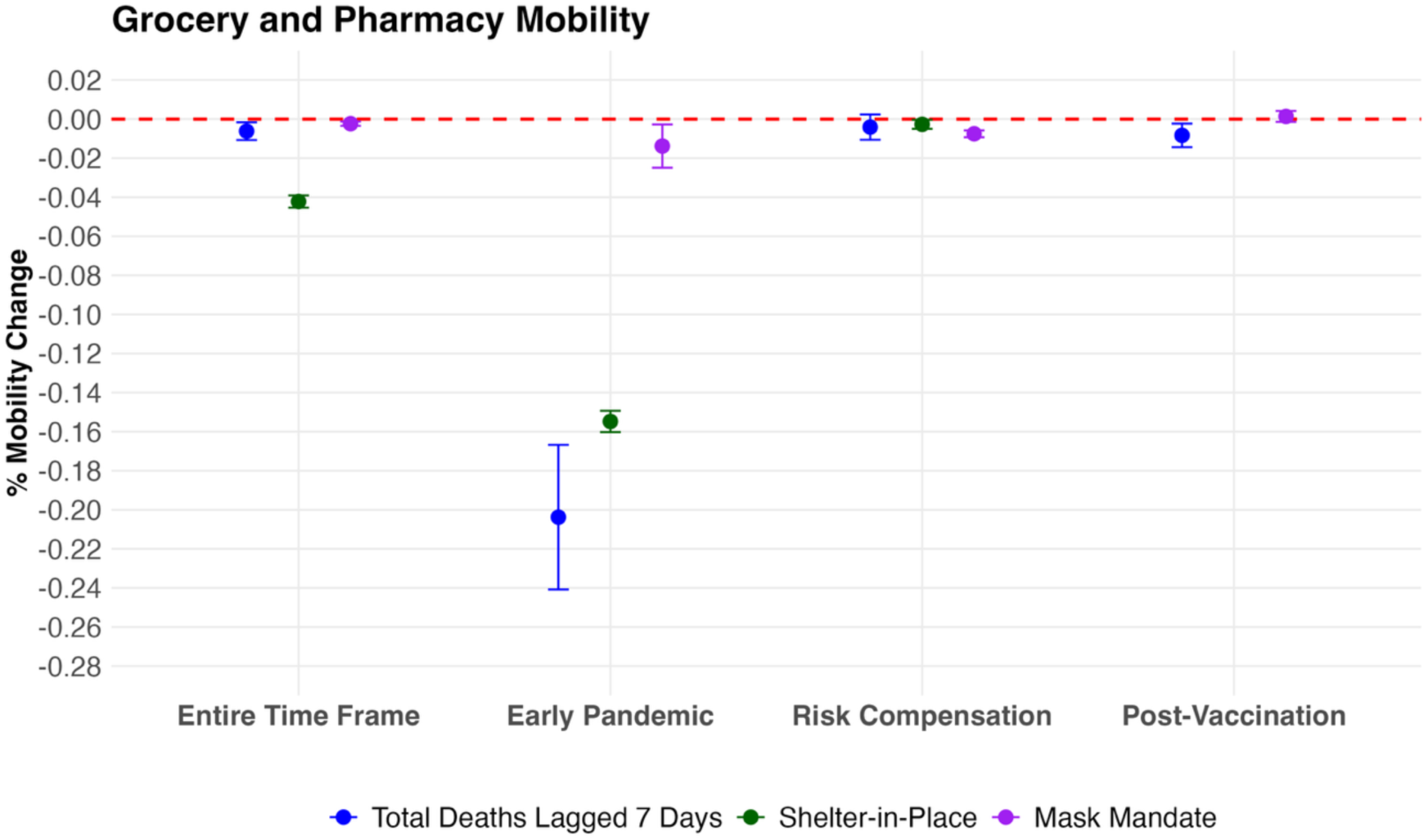
Estimated Effects of Mortality-Driven Risk Perception and Public Health Policies on **Grocery and Pharmacy** Mobility Across Pandemic Phases

**Figure A7.**
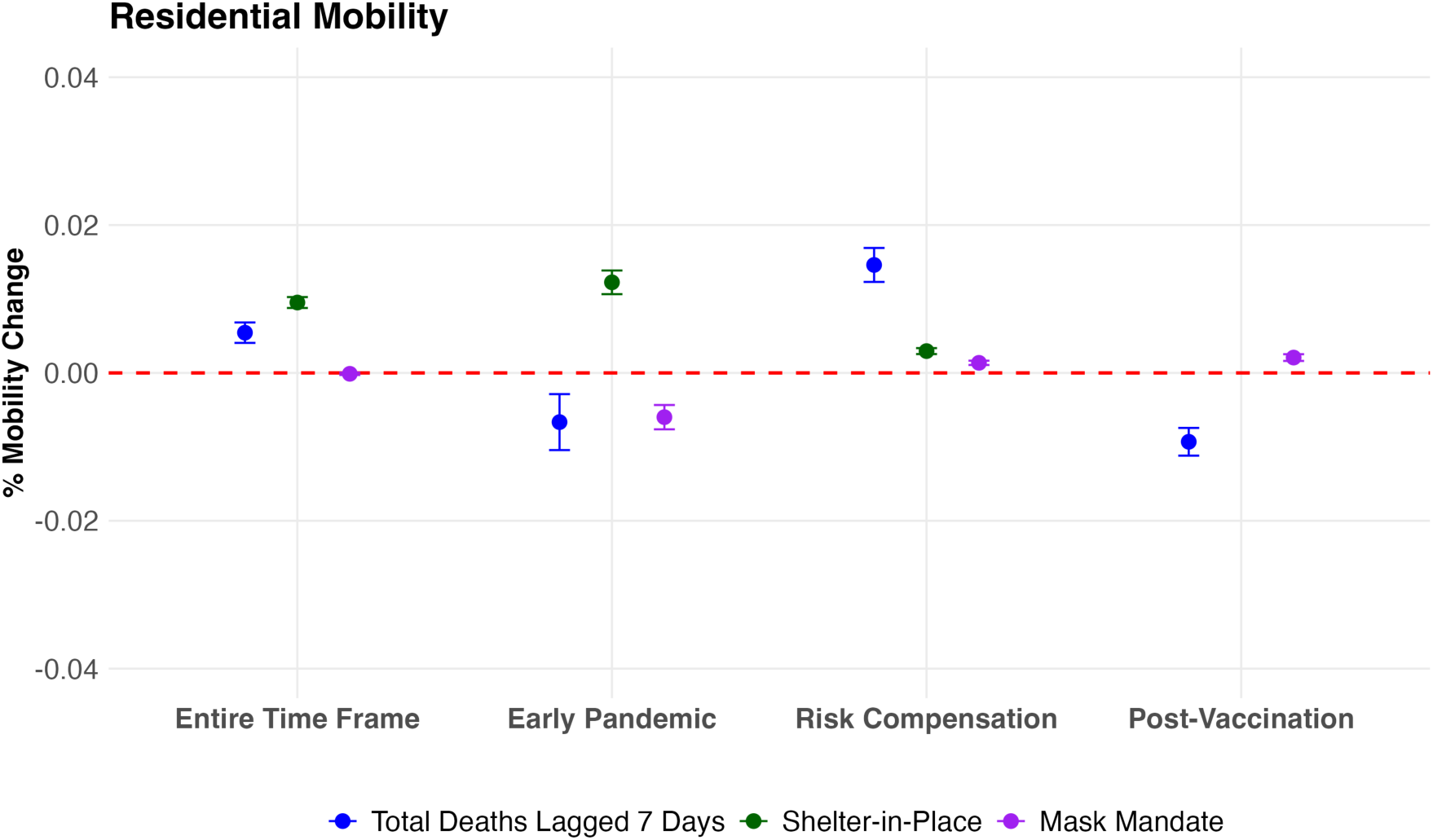
Estimated Effects of Mortality-Driven Risk Perception and Public Health Policies on **Residential** Mobility Across Pandemic Phases

**Figure A8.**
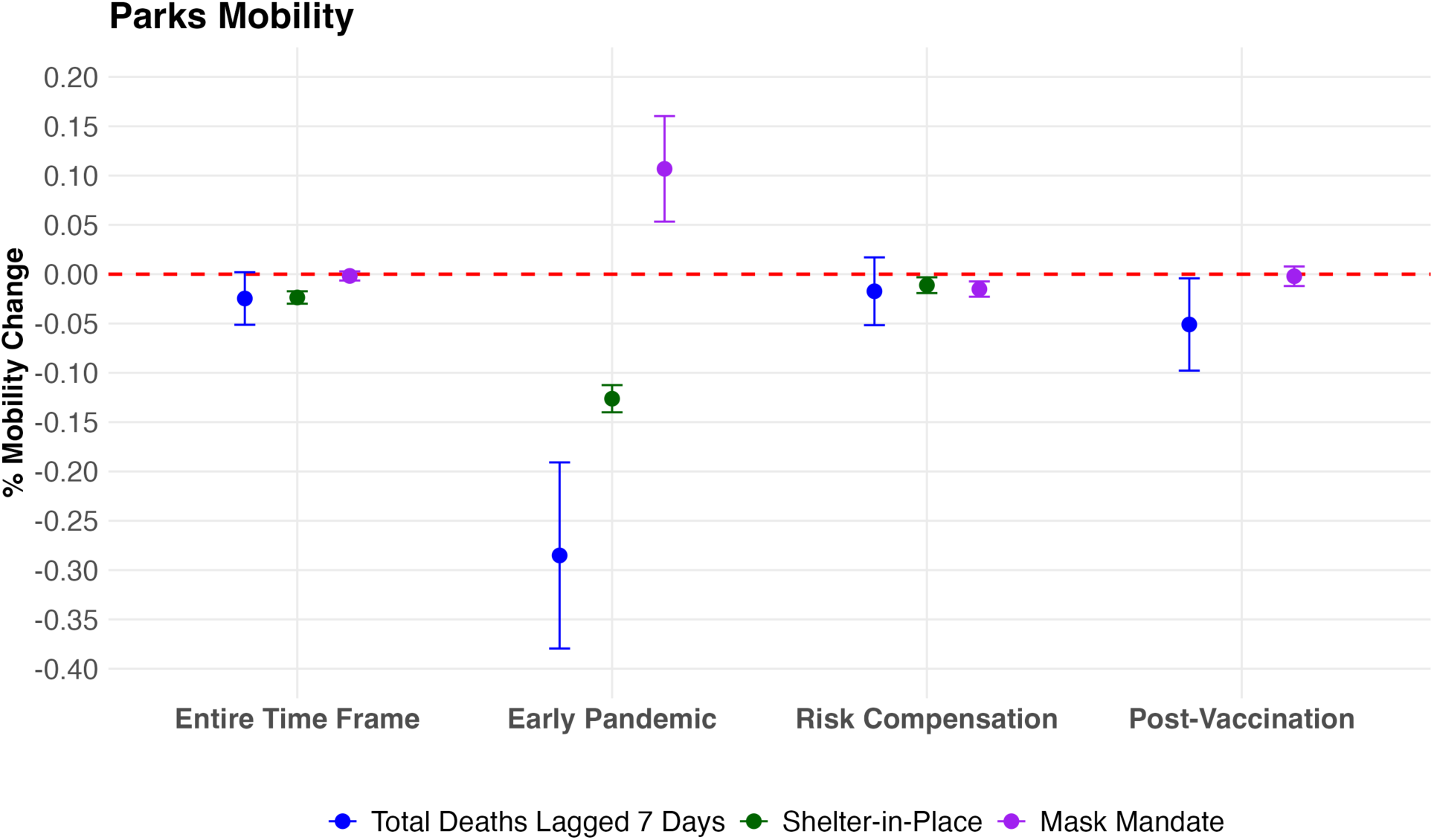
Estimated Effects of Mortality-Driven Risk Perception and Public Health Policies on **Parks** Mobility Across Pandemic Phases

## Footnotes

1 For all log-linear regression results, when indicated we report exact percentage changes using the transformation (*e*^*β*^ − 1) × 100, where *β* is estimated coefficient. This conversion provides an accurate interpretation of the effect size in percentage terms.

